# Pre-existing cardiovascular disease rather than cardiovascular risk factors drives mortality in COVID-19

**DOI:** 10.1101/2020.12.02.20242933

**Authors:** Kevin O’Gallagher, Anthony Shek, Daniel M. Bean, Rebecca Bendayan, James T. H. Teo, Richard J. B. Dobson, Ajay M. Shah, Rosita Zakeri

**Affiliations:** King’s College London British Heart Foundation Centre, School of Cardiovascular Medicine & Sciences, London, UK; Department of Biostatistics and Health Informatics, Institute of Psychiatry, Psychology and Neuroscience, King’s College London, UK; NIHR Biomedical Research Centre at South London and Maudsley NHS Foundation Trust and King’s College London, UK; King’s College Hospital NHS Foundation Trust, London, UK; Health Data Research UK London, Institute of Health Informatics, University College London, UK

**Keywords:** COVID-19, cardiovascular disease, cardiovascular risk factors, hypertension, diabetes

## Abstract

**Background:** The association between cardiovascular (CV) risk factors, such as hypertension and diabetes, established CV disease (CVD), and susceptibility to CV complications or mortality in COVID-19 remains unclear.

**Methods:** We conducted a cohort study of consecutive adults hospitalised for severe COVID-19 between 1^st^ March and 30^th^ June 2020. Pre-existing CVD, CV risk factors and associations with mortality and CV complications were ascertained.

**Findings:** Among 1,721 patients (median age 71 years, 57% male), 349 (20.3%) had pre-existing CVD (CVD), 888 (51.6%) had CV risk factors without CVD (RF-CVD), 484 (28.1%) had neither. Patients with CVD were older with a higher burden of non-CV comorbidities. During follow-up, 438 (25.5%) patients died: 37% with CVD, 25.7% with RF-CVD and 16.5% with neither. CVD was independently associated with in-hospital mortality among patients <70 years of age (adjusted HR 2.43 [95%CI 1.16-5.07]), but not in those ≥70 years (aHR 1.14 [95%CI 0.77-1.69]). RF-CVD were not independently associated with mortality in either age group (<70y aHR 1.21 [95%CI 0.72-2.01], ≥70y aHR 1.07 [95%CI 0.76-1.52]). Most CV complications occurred in patients with CVD (66%) versus RF-CVD (17%) or neither (11%; p<0.001). 213 [12.4%] patients developed venous thromboembolism (VTE). CVD was not an independent predictor of VTE.

**Interpretation:** In patients hospitalised with COVID-19, pre-existing established CVD appears to be a more important contributor to mortality than CV risk factors in the absence of CVD. CVD-related hazard may be mediated, in part, by new CV complications. Optimal care and vigilance for destabilised CVD are essential in this patient group.

## Introduction

The novel coronavirus disease 2019 (COVID-19) has heterogeneous clinical manifestations. Early studies demonstrated that older male patients and individuals with long-term health conditions were at highest risk for severe and fatal outcomes^1-3^. Accordingly, cardiovascular (CV) risk factors such as hypertension and diabetes, and chronic CV diseases (CVD), including ischaemic heart disease and heart failure, are highly prevalent among patients admitted to hospital with severe COVID-19. Whilst some individuals with CV risk factors have concomitant established CVD, many do not. In population-based studies, diabetes and chronic CVD, but not hypertension, have been associated with higher mortality^4,5^. At present, patients with either established CVD or CV risk factors are considered to be vulnerable individuals^6^. However, it remains unclear whether an increased susceptibility to severe COVID-19 in patients with CV risk factors is driven by co-existent CVD, or whether patients with CV risk factors without established CVD have a similarly severe course.

The relationship between CVD and COVID-19 may also be bidirectional. The severe acute respiratory syndrome coronavirus-2 (SARS-CoV-2) is reported to directly infect the endothelium and possibly the heart^7,8^, which could precipitate CV complications. Isolated case reports of fulminant myocarditis or pericarditis have been attributed to COVID-19^9-11^, although the incidence and mechanism of such complications is debated. Furthermore, while patients with pre-existing CVD may be at increased risk of CV complications^12,13^, it is not clear the extent to which these represent recurrent or decompensated CVD rather than *de novo* complications, nor whether the risk also applies to patients with CV risk factors.

To address these questions, we evaluated outcomes associated with pre-existing CVD and CV risk factors, in a large multi-ethnic cohort of patients hospitalised for severe COVID-19. Our aims were to determine a) the relative risks of in-hospital mortality and CV complications for individuals with COVID-19 and pre-existing CVD versus CV risk factors without established CVD, and b) factors associated with the occurrence of major CV complications in patients with COVID-19.

## Methods

### Approvals

This study was conducted under London South East Research Ethics Committee approval (reference 12/LO/2048) granted to the King’s Electronic Records Research Interface (KERRI); specific work on COVID-19 was reviewed with expert patient input on a virtual committee with Caldicott Guardian oversight.

### Study Design

We conducted a retrospective observational cohort study of consecutive adult patients (age >18 years) admitted with COVID-19 to King’s College Hospital NHS Foundation Trust (comprising two hospitals: King’s College Hospital and Princess Royal University Hospital), between 1^st^ March and 30^th^ June 2020. All patients had a positive RT-PCR antigen test for SARS-CoV-2. Only patients admitted to hospital for ≥24 hours were included. A subset of this cohort has been reported previously^14,15^.

### Data sources and processing

Structured and unstructured data were extracted from the electronic health record (EHR) using previously described natural language processing (NLP) informatics tools belonging to the CogStack ecosystem^16^, DrugPipeline^17^, MedCAT^18^, and MedCATtrainer^19^. Clinician case review was used for additional validation (**Supplemental Methods**).

### Exposures and outcomes

CV risk factors were defined as a recorded clinical diagnosis of hypertension, diabetes mellitus, or self-reported current or past smoking status, in the absence of documented CVD. Pre-existing established CVD was defined as a previous record of ≥1 of the following diagnoses: myocardial infarction (MI), heart failure, myocarditis, pericarditis, endocarditis, atrial or ventricular arrhythmia, and valvular heart disease (defined as severe^20^). Additional details are provided in the **Supplemental Methods**. CV risk factors and pre-existing CVD were categorised as present if they had been recorded in the EHR at any time up to the day of admission (or including the day of admission, when recorded as a pre-existing condition). Data were also collected for age, sex, ethnicity, body mass index (BMI), non-CV comorbidities (asthma, chronic obstructive pulmonary disease [COPD], chronic kidney disease [CKD]), previous venous thromboembolism [VTE] comprising deep vein thrombosis [DVT] or pulmonary embolism [PE]) and clinical examination and routinely collected blood results on admission. BMI categories were defined as underweight (<18.5 kg/m^2^), normal (18.5–24.9 kg/m^2^), overweight (25–29.9 kg/m^2^), and obese (≥30kg/m^2^), and the most recent value used (median day of admission [IQR −1 to +5 days]). CV drug therapy at the time of admission was recorded including Angiotensin-Converting Enzyme Inhibitors (ACEI), Angiotensin Receptor Blockers (ARB), aldosterone antagonists, beta-blockers, calcium channel antagonists, loop diuretic agents, statins, antiplatelet and anticoagulant therapy. High sensitivity cardiac troponin T (hs-cTnT) plasma levels were defined as normal when below the 99th percentile of normal values, i.e. 14 ng/L.

The primary outcome was in-hospital mortality. Secondary outcomes included any CV complication related to COVID-19 and incident VTE. A CV complication was defined as a new CVD diagnosis or decompensation of pre-existing CVD (additional details provided in the **Supplemental Methods**), recorded in the EHR on presentation to hospital or at any time during admission. Hospital admission date was used as the start of follow-up. Outcomes were ascertained through to death, discharge, or 31^st^ July 2020, whichever was earlier.

### Statistical analyses

Categorical variables are reported as frequency (%), continuous variables as mean (SD) or median (LQ-UQ), as appropriate. Patient characteristics were compared across 3 groups: patients with pre-existing established CVD (CVD), CV risk factors without CVD (RF-CVD), and no CVD or CV risk factors, using the Chi-squared goodness of fit or Fisher’s exact test (categorical variables), one-way analysis of variance (continuous variables) or Kruskal-Wallis/Wilcoxon rank-sum tests (for non-normally distributed data). The Bonferroni correction was used for individual comparisons. Missing data for blood results were imputed using the multiple imputation approach by chain equations^21^. Imputed variables had less than 25% missing data.

Cumulative incidence plots displaying the probability of in-hospital mortality and discharge were constructed based on a competing risks analysis. To evaluate the association between patient group and mortality, we used Cox proportional hazards regression, with admission date as the start of follow-up and in-hospital mortality as the dependent variable. Unadjusted models examined the effect of patient group, subsequent models were incrementally adjusted for i) demographic variables: age, sex, ethnicity, and, ii) non-CV comorbidities and medications on admission (fully adjusted model). Age was modelled as a categorical variable to allow for potentially non-linear association with outcomes (<40, 40-49, 50-59, 60-69, 70-79, 80+ years). Comorbidities were modelled as binary variables. Patients without pre-existing CVD or CV risk factors were used as the reference group. The proportional hazard assumption was examined graphically and using formal tests, as described by Grambsch^22^; no major deviations from this assumption were observed.

To investigate the association between CV complications and prognosis we performed logistic regression models with patient groups stratified by the presence or absence of CV complications as an independent categorical variable. For secondary outcomes of CV complications or VTE, logistic regression models were constructed with i) patient group and ii) individual CV risk factors or CVDs as binary predictor variables. Unadjusted, demographic-adjusted, and fully adjusted regression models were performed as described above. When individual CVDs were examined, myocarditis and pericarditis were not included in the models, due to their low prevalence. As BMI was missing in >30% of patients, primary analyses were performed without adjustment for BMI. Sensitivity analyses were performed i) restricted to patients with BMI data available including adjustment for BMI as a continuous variable (fully adjusted model), and ii) in the subset of individuals who were discharged or died (i.e. excluding patients still in hospital). Analyses were performed using STATA/IC (v16.1; StataCorp LLC, TX).

## Results

### Study population

Between 1^st^ March and 30^th^ June 2020, 1,721 patients were admitted with COVID-19 (median age 71 years [IQR 56-83], 56.6% male). Of these, 349 (20.3%) patients had CVD, 888 (51.6%) patients had RF-CVD, and 484 (28.1%) patients had neither. Patients with CVD were older than patients with RF-CVD or neither but had a similar distribution of sex (**Table 1**).

**Table 1.**
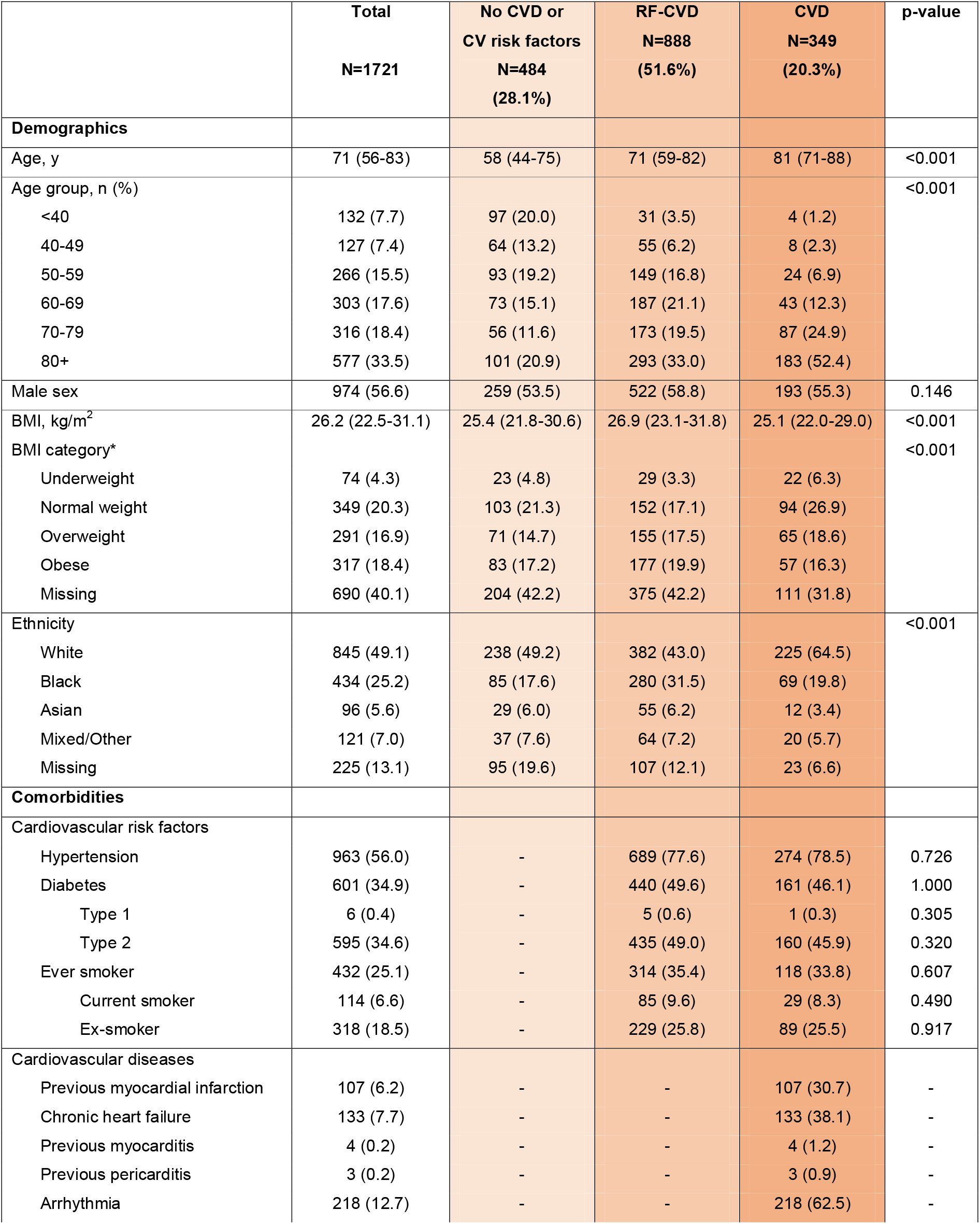
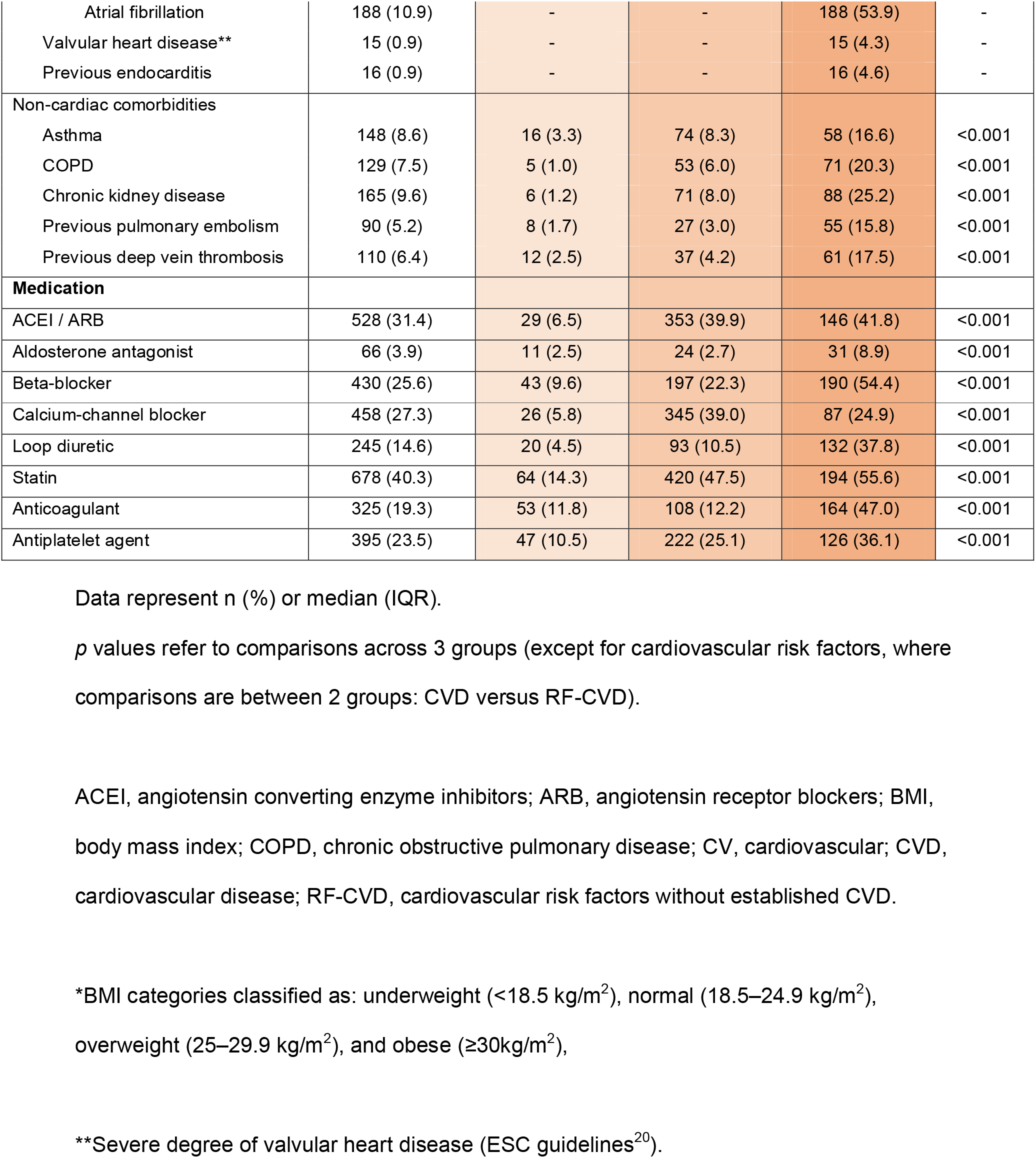
Patient characteristics.

|  | **Total**<br>**N=1721** | **No CVD or**<br>**CV risk factors**<br>**N=484**<br>**(28.1%)** | **RF-CVD**<br>**N=888**<br>**(51.6%)** | **CVD**<br>**N=349**<br>**(20.3%)** | **p-value** |
| --- | --- | --- | --- | --- | --- |
| **Demographics** |  |  |  |  |  |
| Age, y | 71 (56-83) | 58 (44-75) | 71 (59-82) | 81 (71-88) | <0.001 |
| Age group, n (%) |  |  |  |  | <0.001 |
| <40 | 132 (7.7) | 97 (20.0) | 31 (3.5) | 4 (1.2) |  |
| 40-49 | 127 (7.4) | 64 (13.2) | 55 (6.2) | 8 (2.3) |  |
| 50-59 | 266 (15.5) | 93 (19.2) | 149 (16.8) | 24 (6.9) |  |
| 60-69 | 303 (17.6) | 73 (15.1) | 187 (21.1) | 43 (12.3) |  |
| 70-79 | 316 (18.4) | 56 (11.6) | 173 (19.5) | 87 (24.9) |  |
| 80+ | 577 (33.5) | 101 (20.9) | 293 (33.0) | 183 (52.4) |  |
| Male sex | 974 (56.6) | 259 (53.5) | 522 (58.8) | 193 (55.3) | 0.146 |
| BMI, kg/m<sup>2</sup> | 26.2 (22.5-31.1) | 25.4 (21.8-30.6) | 26.9 (23.1-31.8) | 25.1 (22.0-29.0) | <0.001 |
| BMI category* |  |  |  |  | <0.001 |
| Underweight | 74 (4.3) | 23 (4.8) | 29 (3.3) | 22 (6.3) |  |
| Normal weight | 349 (20.3) | 103 (21.3) | 152 (17.1) | 94 (26.9) |  |
| Overweight | 291 (16.9) | 71 (14.7) | 155 (17.5) | 65 (18.6) |  |
| Obese | 317 (18.4) | 83 (17.2) | 177 (19.9) | 57 (16.3) |  |
| Missing | 690 (40.1) | 204 (42.2) | 375 (42.2) | 111 (31.8) |  |
| Ethnicity |  |  |  |  | <0.001 |
| White | 845 (49.1) | 238 (49.2) | 382 (43.0) | 225 (64.5) |  |
| Black | 434 (25.2) | 85 (17.6) | 280 (31.5) | 69 (19.8) |  |
| Asian | 96 (5.6) | 29 (6.0) | 55 (6.2) | 12 (3.4) |  |
| Mixed/Other | 121 (7.0) | 37 (7.6) | 64 (7.2) | 20 (5.7) |  |
| Missing | 225 (13.1) | 95 (19.6) | 107 (12.1) | 23 (6.6) |  |
| **Comorbidities** |  |  |  |  |  |
| Cardiovascular risk factors |  |  |  |  |  |
| Hypertension | 963 (56.0) | - | 689 (77.6) | 274 (78.5) | 0.726 |
| Diabetes | 601 (34.9) | - | 440 (49.6) | 161 (46.1) | 1.000 |
| Type 1 | 6 (0.4) | - | 5 (0.6) | 1 (0.3) | 0.305 |
| Type 2 | 595 (34.6) | - | 435 (49.0) | 160 (45.9) | 0.320 |
| Ever smoker | 432 (25.1) | - | 314 (35.4) | 118 (33.8) | 0.607 |
| Current smoker | 114 (6.6) | - | 85 (9.6) | 29 (8.3) | 0.490 |
| Ex-smoker | 318 (18.5) | - | 229 (25.8) | 89 (25.5) | 0.917 |
| Cardiovascular diseases |  |  |  |  |  |
| Previous myocardial infarction | 107 (6.2) | - | - | 107 (30.7) | - |
| Chronic heart failure | 133 (7.7) | - | - | 133 (38.1) | - |
| Previous myocarditis | 4 (0.2) | - | - | 4 (1.2) | - |
| Previous pericarditis | 3 (0.2) | - | - | 3 (0.9) | - |
| Arrhythmia | 218 (12.7) | - | - | 218 (62.5) | - |
| Atrial fibrillation | 188 (10.9) | - | - | 188 (53.9) | - |
| Valvular heart disease** | 15 (0.9) | - | - | 15 (4.3) | - |
| Previous endocarditis | 16 (0.9) | - | - | 16 (4.6) | - |
| Non-cardiac comorbidities |  |  |  |  |  |
| Asthma | 148 (8.6) | 16 (3.3) | 74 (8.3) | 58 (16.6) | <0.001 |
| COPD | 129 (7.5) | 5 (1.0) | 53 (6.0) | 71 (20.3) | <0.001 |
| Chronic kidney disease | 165 (9.6) | 6 (1.2) | 71 (8.0) | 88 (25.2) | <0.001 |
| Previous pulmonary embolism | 90 (5.2) | 8 (1.7) | 27 (3.0) | 55 (15.8) | <0.001 |
| Previous deep vein thrombosis | 110 (6.4) | 12 (2.5) | 37 (4.2) | 61 (17.5) | <0.001 |
| **Medication** |  |  |  |  |  |
| ACEI / ARB | 528 (31.4) | 29 (6.5) | 353 (39.9) | 146 (41.8) | <0.001 |
| Aldosterone antagonist | 66 (3.9) | 11 (2.5) | 24 (2.7) | 31 (8.9) | <0.001 |
| Beta-blocker | 430 (25.6) | 43 (9.6) | 197 (22.3) | 190 (54.4) | <0.001 |
| Calcium-channel blocker | 458 (27.3) | 26 (5.8) | 345 (39.0) | 87 (24.9) | <0.001 |
| Loop diuretic | 245 (14.6) | 20 (4.5) | 93 (10.5) | 132 (37.8) | <0.001 |
| Statin | 678 (40.3) | 64 (14.3) | 420 (47.5) | 194 (55.6) | <0.001 |
| Anticoagulant | 325 (19.3) | 53 (11.8) | 108 (12.2) | 164 (47.0) | <0.001 |
| Antiplatelet agent | 395 (23.5) | 47 (10.5) | 222 (25.1) | 126 (36.1) | <0.001 |

Data represent n (%) or median (IQR).
*p* values refer to comparisons across 3 groups (except for cardiovascular risk factors, where comparisons are between 2 groups: CVD versus RF-CVD).
ACEI, angiotensin converting enzyme inhibitors; ARB, angiotensin receptor blockers; BMI, body mass index; COPD, chronic obstructive pulmonary disease; CV, cardiovascular; CVD, cardiovascular disease; RF-CVD, cardiovascular risk factors without established CVD.
\*BMI categories classified as: underweight (<18.5 kg/m<sup>2</sup>), normal (18.5–24.9 kg/m<sup>2</sup>), overweight (25–29.9 kg/m<sup>2</sup>), and obese (≥30kg/m<sup>2</sup>),
\*\*Severe degree of valvular heart disease (ESC guidelines<sup>20</sup>).

CVD was more prevalent with increasing age, while RF-CVD was most common between 50-70 years (**Supplemental Figure 1A**). Individuals from non-White ethnic groups had a higher prevalence of RF-CVD whereas CVD was more prevalent in the White group (**Supplemental Figure 1B**). The most frequent CVD diagnoses were arrhythmia (86.2% atrial fibrillation), heart failure, and previous MI, respectively (**Table 1**). 119 (34.1%) patients with CVD had more than one CVD diagnosis. Rates of non-cardiovascular comorbidities were highest in patients with CVD, followed by RF-CVD, versus patients with neither (**Table 1**).

On admission, 83% of patients with hypertension were taking an antihypertensive agent, and 73% of patients with atrial fibrillation were on oral anticoagulation. Rates of ACEI or ARB and betablocker use in heart failure patients were 47% and 62% respectively. In patients with a previous MI, rates of antiplatelet, beta-blocker and statin use were 68%, 64% and 65% respectively.

### Clinical presentation

Physiological parameters and blood biomarkers are displayed in **Table 2**. There were few clinically significant differences in physiological observations between groups, with the exception of a higher mean systolic blood pressure in patients with RF-CVD (**Table 2**).

**Table 2.** Clinical observations and laboratory results.

|  | **Total**<br>**N=1721** | **No CVD or**<br>**CV risk factors**<br>**N=484**<br>**(28.1%)** | **RF-CVD**<br>**N=888**<br>**(51.6%)** | **CVD**<br>**N=349**<br>**(20.3%)** | **p-value** |
| --- | --- | --- | --- | --- | --- |
| **Observations** |  |  |  |  |  |
| Temperature, °C | 37.0 ± 0.02 | 37.1 ± 0.04 | 37.1 ± 0.02 | 36.8 ± 0.03 | <0.001 |
| Heart rate, /min | 85.7 ± 0.7 | 88.3 ± 2.2 | 84.9 ± 0.6 | 83.9 ± 0.9 | 0.058 |
| Systolic blood pressure, mmHg | 127.0 ± 0.7 | 124.2 ± 1.6 | 129.1 ± 0.8 | 125.4 ± 1.3 | 0.003 |
| Diastolic blood pressure, mmHg | 72.6 ± 0.5 | 72.7 ± 0.8 | 73.7 ± 0.9 | 69.8 ± 0.8 | 0.017 |
| Respiratory rate, /min | 20.5 ± 0.1 | 20.4 ± 0.3 | 20.8 ± 0.2 | 20.0 ± 0.3 | 0.085 |
| Oxygen saturation, % | 96.0 ± 0.1 | 96.0 ± 0.3 | 96.0 ± 0.1 | 96.2 ± 0.1 | 0.649 |
| **Laboratory results** |  |  |  |  |  |
| CRP, mg/L | 97.4 ± 2.2 | 87.4 ± 4.3 | 107.9 ± 84.6 | 84.6 ± 4.3 | <0.001 |
| White cell count, x10<sup>9</sup>/L | 9.0 ± 0.2 | 9.5 ± 0.5 | 8.6 ± 0.2 | 9.2 ± 0.4 | 0.130 |
| Lymphocytes, x10<sup>9</sup>/L | 1.4 ± 0.1 | 1.6 ± 0.2 | 1.3 ± 0.2 | 1.3 ± 0.2 | 0.438 |
| Neutrophils, x10<sup>9</sup>/L | 6.9 ± 0.1 | 7.2 ± 0.3 | 6.7 ± 0.1 | 7.1 ± 0.3 | 0.196 |
| Platelets, x10<sup>9</sup>/L | 234.6 ± 2.5 | 238.2 ± 5.1 | 234.3 ± 3.3 | 230.4 ± 5.8 | 0.571 |
| Sodium, mmol/L | 137.8 ± 0.2 | 138.0 ± 0.3 | 137.7 ± 0.2 | 137.7 ± 0.4 | 0.659 |
| Urea, mmol/L | 9.7 ± 0.2 | 6.6 ± 0.3 | 10.6 ± 0.3 | 11.7 ± 0.4 | <0.001 |
| eGFR, mL/min/m<sup>2</sup> | 60.1 ± 0.6 | 72.1 ± 1.0 | 57.3 ± 0.9 | 50.7 ± 1.4 | <0.001 |
| Albumin, g/L | 36.9 ± 0.1 | 38.0 ± 36.7 | 36.7 ± 0.2 | 35.6 ± 0.3 | <0.001 |
| ALP | 99.4 ± 2.2 | 96.1 ± 4.3 | 94.3 ± 3.0 | 117.3 ± 5.0 | <0.001 |
| Bilirubin | 12.3 ± 0.6 | 14.4 ± 1.7 | 10.9 ± 0.5 | 12.9 ± 0.9 | 0.031 |
| High sensitivity cardiac troponin T, ng/L |  |  |  |  |  |
| Baseline* | 21 (9-50) | 11 (5-31) | 22 (11-50) | 43 (19-76) | <0.001 |
| N | 552 | 145 | 308 | 99 |  |
| N (%) elevated** | 349 (63.2) | 63 (43.4) | 203 (65.9) | 83 (83.8) |  |
| Peak | 36 (16-85) | 27 (10-71) | 35 (16-84) | 48 (23-93) | <0.001 |
| N | 742 | 186 | 409 | 147 |  |
| N (%) elevated** | 567 (76.4) | 123 (66.1) | 311 (76.0) | 133 (90.5) |  |

Data reported as mean ± SEM (median [IQR] for high sensitivity cardiac troponin T)
CRP, C-reactive protein; CV, cardiovascular; CVD, cardiovascular disease; eGFR, estimated glomerular filtration rate; ALP, alkaline phosphatase; RF-CVD, cardiovascular risk factors without established CVD.
\*Within 24 hours of admission
\*\*Greater than 14ng/L

Among blood biomarkers, C-reactive protein values were highest in patients with RF-CVD, but similar between patients with CVD and the group with neither CVD nor risk factors (**Table 2**). Renal function was progressively worse across groups, with the lowest eGFR in patients with CVD. Liver function markers were also elevated in patients with CVD versus other groups but remained, on average, within a normal range.

Overall, 742 (43.1%) patients had at least one hs-cTnT measurement, and 552 (32.1%) had a measurement within 24 hours of admission. Patients who did not have a hs-cTnT measurement were older (74.0 vs. 67 years; p<0.001) and more likely to be women (48.4% vs. 36.8%; p<0.001). Among patients with at least one hs-cTnT measurement, elevated values (>14ng/L) were observed in 133/147 (90.5%) patients with CVD, 311/409 (76.0%) patients with RF-CVD and 123/186 (66.1%) patients with no CVD or CV risk factors. Values greater than 10x the upper limit of normal were observed in 23 (15.7%) patients with CVD, 66 (16.1%) patients with RF-CVD, and 22 (11.8%) patients with no CVD or risk factors.

### In-hospital mortality

In-hospital outcomes are displayed in **Table 3**. Overall, 438 (25.5%) patients died, 1,246 (72.4%) were discharged and 37 (2.1%) remained in hospital at the end of the study period. The median length of hospitalisation for patients discharged alive was 9 (IQR 4-17) days and was longer for patients with CVD than the groups without CVD (11 [IQR 5-19] vs 7 [IQR 3-16] days, p<0.001).

**Table 3.** Complications and in-hospital outcomes of patients with COVID-19.

|  | **Total**<br>**N=1721** | **No CVD or**<br>**CV risk factors**<br>**N=484 (28.1%)** | **RF-CVD**<br>**N=888 (51.6%)** | **CVD**<br>**N=349 (20.3%)** | **p-value** |
| --- | --- | --- | --- | --- | --- |
| **Complications** |  |  |  |  |  |
| Cardiac |  |  |  |  |  |
| Acute myocardial infarction | 68 (4.0) | 4 (0.8) | 21 (2.4) | 43 (12.3) | <0.001 |
| Acute heart failure | 151 (8.8) | 12 (2.5) | 43 (4.8) | 96 (27.5) | <0.001 |
| Myocarditis | 12 (0.7) | 0 | 9 (1.0) | 3 (0.9) | 0.090 |
| Pericarditis | 2 (0.1) | 1 (0.2) | 1 (0.1) | 0 | 0.688 |
| Arrhythmia* | 314 (18.3) | 40 (8.3) | 99 (11.2) | 175 (50.1) | <0.001 |
| Atrial fibrillation | 266 (15.5) | 28 (5.8) | 74 (8.3) | 164 (47.0) | <0.001 |
| Number of cardiac complications** |  |  |  |  | <0.001 |
| 0 | 1,290 (75.0) | 433 (89.5) | 738 (83.1) | 119 (34.1) |  |
| 1 | 325 (18.9) | 45 (9.3) | 129 (14.5) | 151 (43.3) |  |
| 2+ | 106 (6.2) | 6 (1.2) | 21 (2.4) | 79 (22.6) |  |
| Venous thromboembolism |  |  |  |  |  |
| Pulmonary embolism | 151 (8.8) | 41 (8.5) | 66 (7.4) | 44 (12.6) | 0.015 |
| Deep vein thrombosis | 98 (5.7) | 21 (4.3) | 43 (4.8) | 34 (9.7) | 0.001 |
| Extra-cardiac |  |  |  |  |  |
| Acute kidney injury*** | 266 (15.5) | 30 (6.2) | 164 (18.5) | 72 (20.6) | <0.001 |
| **Outcomes** |  |  |  |  |  |
| Died in hospital | 438 (25.5) | 80 (16.5) | 228 (25.7) | 130 (37.3) | <0.001 |
| ICU admission | 224 (13.0) | 74 (15.3) | 127 (14.3) | 23 (6.6) | <0.001 |
| Death or ICU admission | 587 (34.1) | 132 (27.3) | 311 (35.0) | 144 (41.3) | <0.001 |
| Discharged from hospital alive | 1,246 (72.4) | 393 (81.2) | 639 (72.0) | 214 (61.3) | <0.001 |
| Hospital length of stay<sup>§</sup>, days | 9 (4-17) | 7 (3-16) | 8 (4-18) | 11 (5-19) | <0.001 |

Data presented as n (%) or median (IQR). Table includes all in-hospital diagnoses during the admission (including new and recurrent diagnoses).
\*Any physician-identified cardiac arrhythmia
\*\* Number of physician-diagnosed CV complications from the following: acute myocardial infarction, heart failure, myocarditis, pericarditis, arrhythmia including AF, and endocarditis.
\*\*\* Acute kidney injury was defined according to the Kidney Disease: Improving Global Outcomes definition<sup>36</sup>.
§ Among patients discharged

In-hospital mortality was greatest among patients with CVD (37.3%), intermediate in patients with RF-CVD (25.7%), and lowest among patients with neither (16.5%). **Figure 1** displays cumulative incidence plots of the probability of in-hospital death or discharge over time for each group. For the overall cohort, there was a positive association between CVD and in-hospital mortality in unadjusted (HR 2.17 [95%CI 1.64-2.87], p<0.001), and demographic-adjusted models (adjusted HR 1.45 [95%CI 1.09-1.94], p=0.012). In fully adjusted models additionally accounting for non-CV comorbidities and baseline medications, there was a positive trend (aHR 1.36 [95%CI 0.97-1.92], p=0.076). This effect was principally driven by a prognostic association in patients under 70 years of age (**Figure 2A**), whereas the effect of CVD was smaller and non-statistically significant among patients aged 70 years and older (**Figure 2B**). RF-CVD conferred an increased risk of mortality for the overall cohort in unadjusted analyses (HR 1.51 [95%CI 1.17-1.95], p=0.002), but not in demographic-adjusted (aHR 1.17 [95%CI 0.90-1.53], p=0.233) or fully adjusted models (aHR 1.13 [95%CI 0.85-1.51], p=0.388). RF-CVD were not significantly associated with mortality in patients older or younger than 70 years of age (**Figure 2A-B**).

**Figure 1.**
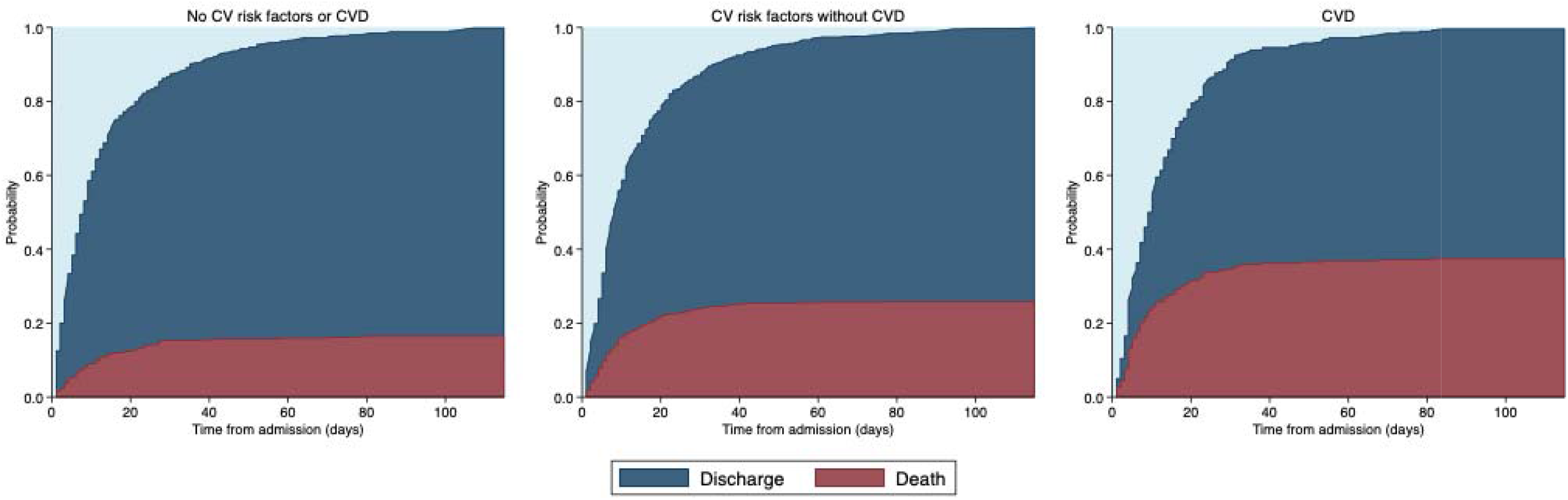
Cumulative incidence plots displaying the probability of in-hospital death and discharge over time. The light blue region represents the probability of being alive and still in hospital at the time of study close. CV, cardiovascular; CVD, cardiovascular disease.

**Figure 2.**
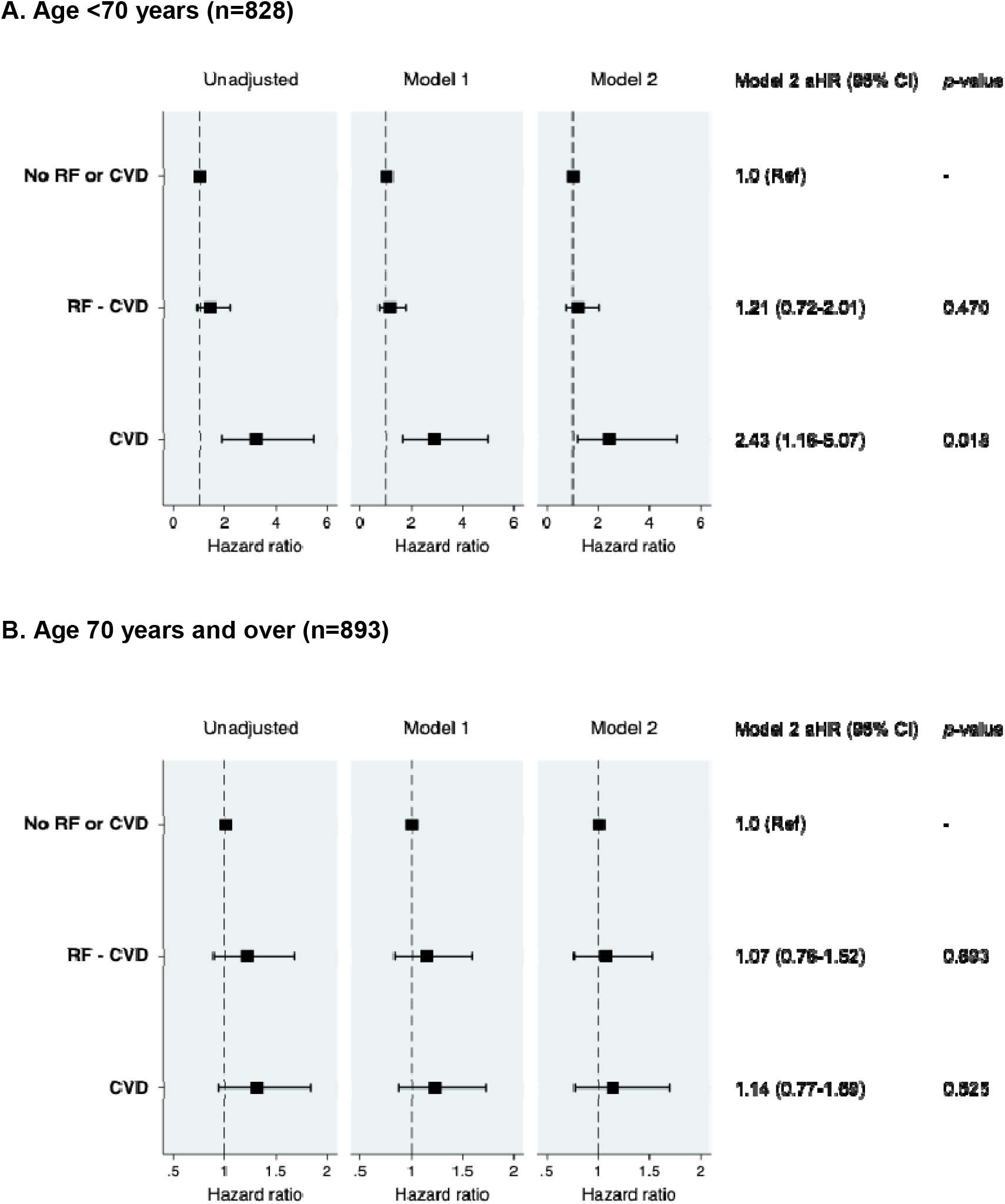
Risk of in-hospital mortality stratified by age and cardiovascular risk group. aHR, adjusted hazard ratio; CVD, cardiovascular disease; RF, cardiovascular risk factors; RF-CVD, cardiovascular risk factors without established CVD. Model 1 adjusted for age, sex and ethnicity. Model 2 adjusted for age, sex, ethnicity, non-cardiac comorbidities (asthma, COPD, chronic renal failure, pulmonary embolism, DVT) and medications (ACEI or ARB, aldosterone receptor antagonists, beta-blockers, calcium channel blockers, loop diuretics, statins, anticoagulants, antiplatelet agents).

The main findings were unchanged in sensitivity analyses additionally adjusting for BMI, in the subset of patients for whom BMI data were available (**Supplemental Figure 2A-B**), or in analyses limited to patients who died or were discharged (i.e. excluding current in-patients; **Supplemental Figure 3A-B**),

### Cardiovascular complications

Cardiovascular complications occurred in 431 (25.0%) patients, with two-thirds occurring in patients with CVD (n=230, 65.9%). Patients with RF-CVD also had a higher CV complication rate than patients with no CVD or CV risk factors (16.9% versus 10.5%, Bonferroni adjusted p<0.001). The most frequent CV complications were cardiac arrhythmias (84.7% atrial fibrillation), followed by acute heart failure (distinct from myocarditis) and acute MI, respectively (**Table 3**). The incidence of clinician-diagnosed myocarditis was low (0.7%). When arrhythmia-related complications were excluded, 59% of complications occurred in patients with CVD, 33% in patients with RF-CVD, and 8% in patients with neither.

In patients with CVD, the majority of CV complications represented exacerbations or decompensation of the underlying CVD, rather than a new presentation, e.g. 86% of myocardial infarctions occurred in individuals with a previous myocardial infarction (**Supplemental Figure 4**). Exacerbation/decompensation of arrhythmia-related complications (including atrial fibrillation), defined as new or worsening symptoms, haemodynamic instability or poor heart rate control, also occurred most frequently in individuals with a pre-existing arrhythmia diagnosis. Among specific CVDs and risk factors, pre-existing AF was associated with the highest adjusted odds of having any CV complication, followed by previous MI (**Figure 3A**). For non-arrhythmia related CV complications, the highest adjusted odds were seen in patients with a previous myocardial infarction (**Figure 3B**).

**Figure 3.**
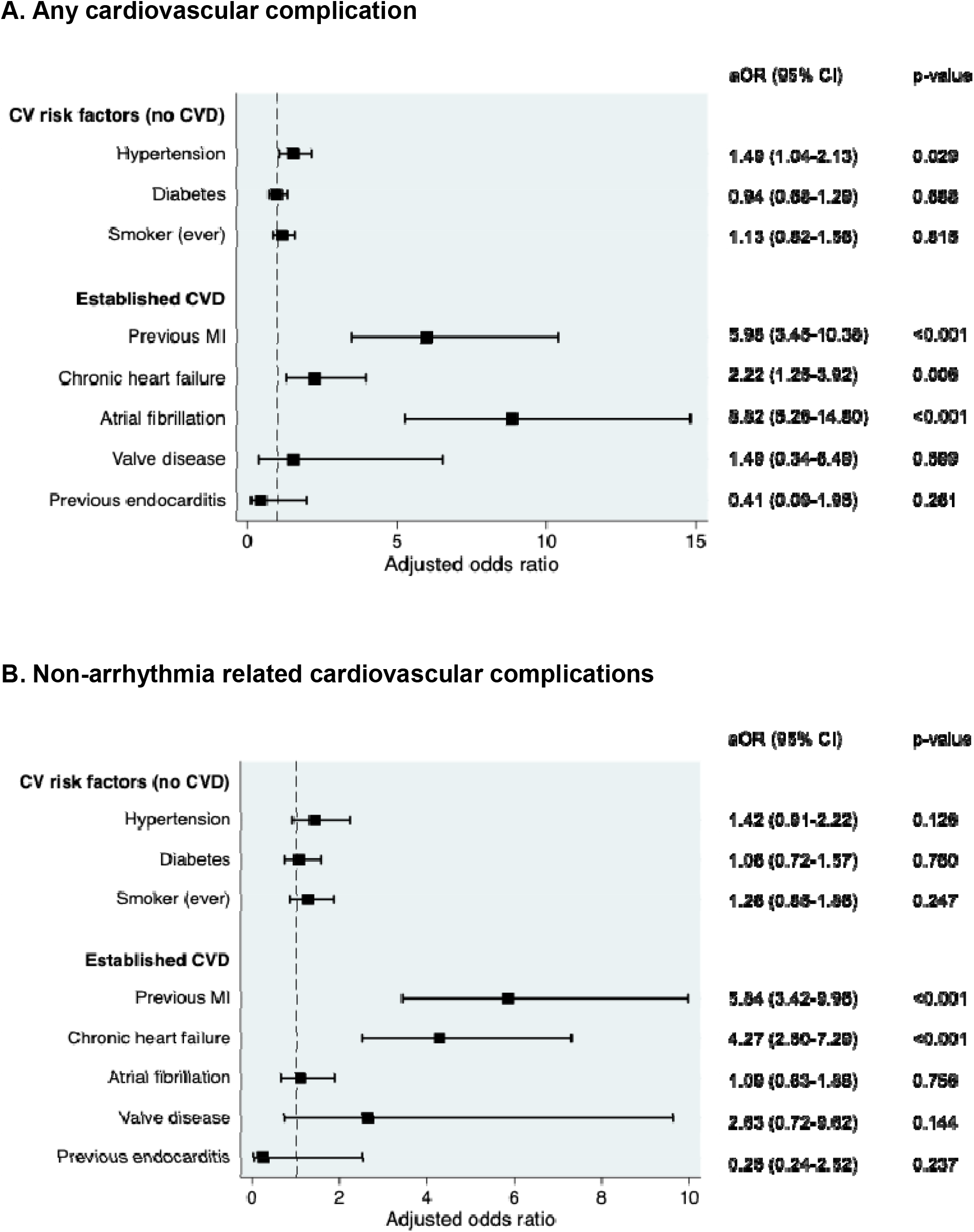
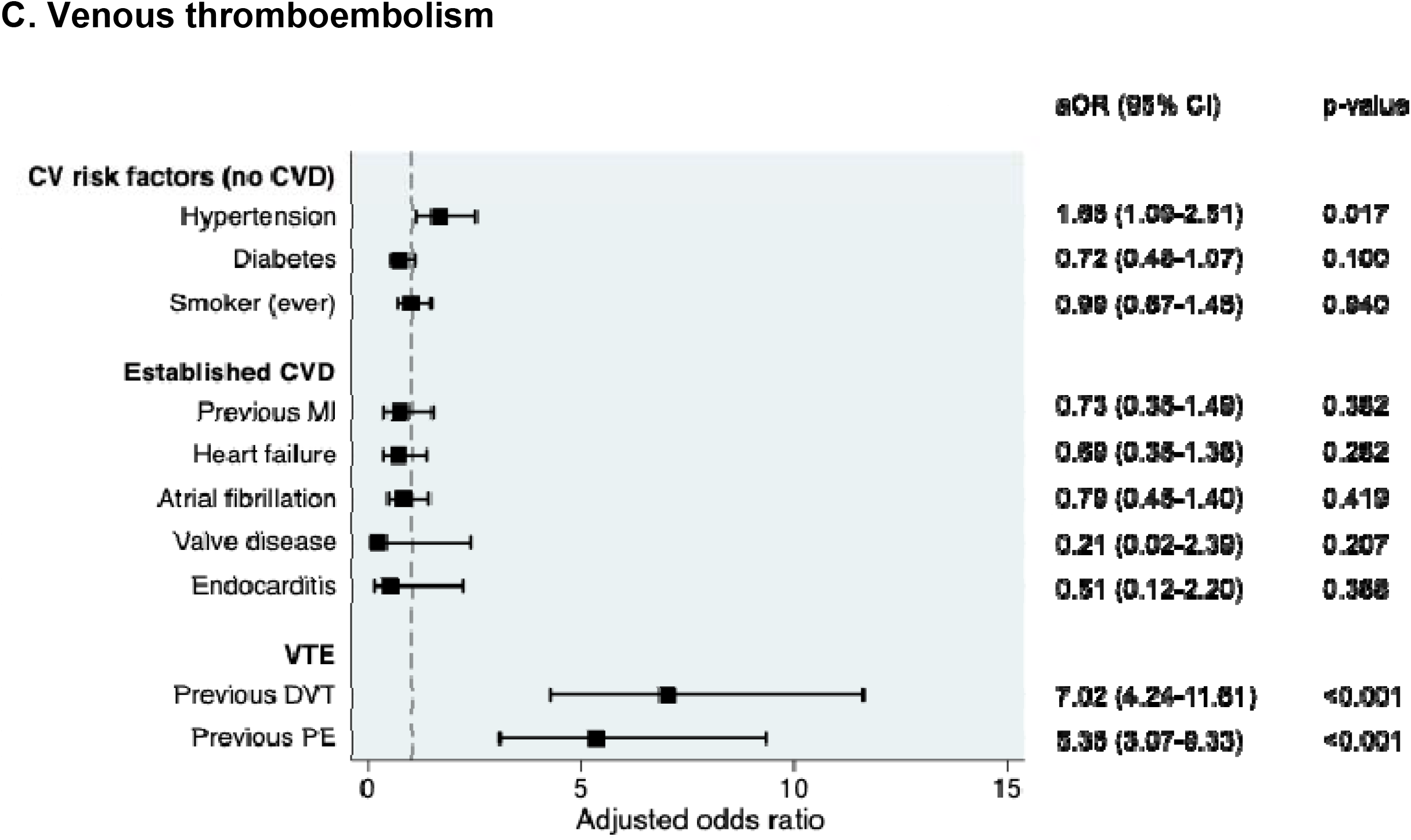
Risk of COVID-19 related complications by cardiovascular risk group. aOR, adjusted odds ratio; CV, cardiovascular; CVD, cardiovascular disease; DVT, deep vein thrombosis; PE, pulmonary embolism; MI, myocardial infarction; VTE, venous thromboembolism. Each model is adjusted for the variables listed, as well as age, sex, ethnicity, non-cardiac comorbidities (asthma, COPD, chronic renal failure, pulmonary embolism, DVT) and medications (ACEI or ARB, aldosterone receptor antagonists, beta-blockers, calcium channel blockers, loop diuretics, statins, anticoagulants, antiplatelet agents).

When CV complications were defined by cardiac biomarker elevation, in the subset of patients with a hs-cTnT measurement (n=742), the presence of any troponin elevation (**Supplemental Figure 5A**) and troponin-elevation greater than 10x normal (**Supplemental Figure 5B**) were both associated with increased odds of in-hospital mortality in all patient groups.

The incidence of VTE was higher in patients with CVD versus RF-CVD or neither (18.3% versus 10.9% and 10.7% respectively, p<0.001 for both comparisons). However, among CVDs and CV risk factors, hypertension (in the absence of established CVD) was the only independent CV predictor of VTE. Patients with previous VTE (n=166, 48% anticoagulated at admission) had the highest rate of new (incident) VTE (49.4% vs 8.4% with no previous VTE p<0.001), and previous VTE was the strongest predictor of incident VTE, including adjustment for baseline anticoagulation use (**Figure 3B**).

## Discussion

Both pre-existing established CVD and CV risk factors such as hypertension and diabetes have been cited as associated with severe and fatal COVID-19 but their relative importance remains unclear. In the present study, we investigated the inter-relationship between CVD, CV risk factors, CV complications and mortality among 1,721 consecutive patients hospitalised due to COVID-19. Overall, 20% of the cohort had CVD while an additional 50% had CV risk factors without yet having developed CVD (RF-CVD). A major finding is that the age- and sex-adjusted mortality risk is markedly increased in patients under the age of 70 years with CVD but is only modestly and non-significantly increased in those with RF-CVD. The mortality risk associated with CVD appears much lower in individuals above 70 years of age. We also found that 1 in 4 patients hospitalised with COVID-19 experienced a CV complication, with cardiac arrhythmias representing the most common diagnosis, and that the majority of CV complications and myocardial injury occurred in patients with CVD. Myocardial injury as indicated by an elevated troponin level was an independent predictor of mortality. Taken together, these findings support the notion that pre-existing established CVD rather than CV risk factors *per se* influence mortality in severe COVID-19 and that this effect may be driven at least in part by CV complications and injury.

### Pre-existing cardiovascular disease

Evidence to date suggests that patients with pre-existing CVD exhibit more severe COVID-19, with a greater likelihood of hospitalisation^15,23,24^. Accordingly, the prevalence of CVD in our study was similar to other large hospital cohorts^23,25^. We corroborate previous reports showing that a history of CVD is associated with greater risk of COVID-19-related mortality^4,23,24^. Interestingly, we identify an interaction with age, wherein the increase in mortality risk is mainly apparent in people below the age of 70 years. The reasons why older individuals do not also manifest higher CVD-related mortality warrant further investigation but may be related to a higher non-CVD-related mortality risk due to factors such as frailty, other comorbidities and immunosenescence, such that CVD has relatively minor additional effects. Similar age-dependent mortality effects have been reported in other studies^4,5^.

### Cardiovascular risk factors

CV risk factors such as hypertension (56%) and diabetes (35%) were more prevalent than established CVD in our in-patient cohort, similar to other UK studies^23^. Patients from non-White minority ethnic groups had especially high rates of CV risk factors. Despite their high prevalence, there are conflicting data regarding the prognostic impact of common CV risk factors in COVID-19. In hospitalized cohorts such as the UK ISARIC study, diabetes had only a marginal independent effect on mortality risk, similar to the findings of the current study^23^. The majority of patients in ISARIC had uncomplicated diabetes while hypertension was not assessed. In the population-based OpenSAFELY study, diabetes was independently associated with a higher mortality, with a HR between 1.31 (1.24-1.37) and 1.95 (1.83-2.08) by HbA1c classification, whereas hypertension was not independently associated with mortality^4^. Another large UK population-based study reported a 1.8-fold higher mortality risk for patients with type 2 diabetes after adjustment for multiple factors including CVD^26^. These apparently divergent findings may be reconciled by considering that mortality rates in population-based studies reflect the risk of infection as well as risk of mortality once infected. For example, the higher mortality risk associated with diabetes in population-based studies was suggested to be related, at least in part, to the level of glycaemic control, which also may impact directly on infection risk^5^. Therefore, while patients with diabetes may be more likely to be admitted, the current study and previous reports^23^ suggest that the mortality risk in this group may largely relate to the co-existence of CVD or other complications.

It should be acknowledged, however, that there may be interactions among CV risk factors and other variables that affect mortality risk. For example, the relationship between diabetes and mortality is reported to be much steeper in non-white patients below the age of 70 years than in other groups^5^. Mortality risks may also be modified by the rates and intensity of medications such as anti-hypertensive treatment^5^. In the current study, rates of medication use were relatively high (e.g. 83% patients with hypertension on treatment), which could also reduce the impact of CV risk factors.

### Cardiovascular complications

A greater mortality risk associated with pre-existing CVD as compared to CV risk factors without CVD raises questions about the potential mechanisms underlying the higher risk. It has been proposed by several authors that endothelial dysfunction may be a major contributor to severe COVID-19^27,28^. Accordingly, pre-existing endothelial dysfunction may increase the likelihood of developing severe endothelial and vascular impairment with COVID-19. However, such a mechanism would not readily explain the differential risk between established CVD and CV risk factors since both conditions are associated with endothelial dysfunction (and the majority of patients with CVD have CV risk factors). An alternative possibility is that patients with pre-existing established CVD are more prone to develop further cardiac injury and dysfunction which, in combination with pulmonary and right heart problems that represent the major manifestations of severe COVID-19, leads to life-threatening illness.

To explore this possibility, we also analysed CV complications in the current study. The use of semi-automated pipelines to capture all clinician-diagnosed CV events documented in the electronic health record minimised selection and indication bias. With this, our data demonstrate a high frequency of CV events overall (25%), with the majority (73%) representing arrhythmias, and rates of atrial fibrillation comparable to smaller studies^24,29,30^. Other complications included problems such as acute MI and acute heart failure whereas few clinically-diagnosed cases of myocarditis were observed (0.7%). Patients with pre-existing CVD had higher rates of CV complications than those with CV risk factors without CVD or patients without either CVD or CV risk factors. They also had higher rates of VTE but, whereas CVD was an independent predictor for the occurrence of CV complications, it was not a predictor of VTE. Importantly, we also found that a high proportion of COVID-19 related CV complications (mainly cardiac) represent exacerbated or destabilised pre-existing CVD, rather than new presentations. This was particularly true for patients with atrial fibrillation (91% destabilised disease), previous MI (86% recurrent MI), and to a lesser extent HF (68% decompensated disease). Taken together, these findings suggest that the detrimental impact of pre-existing CVD on COVID-19 severity and mortality may be mediated mainly by increased cardiac problems rather than systemic vascular abnormalities such as VTE. In support of this idea, we also found that myocardial injury as assessed by troponin elevation was most prevalent in patients with pre-existing CVD and that it was strongly associated with mortality.

The high incidence of cardiac arrhythmias, including atrial fibrillation, observed in this study may have multiple precipitants, such as myocardial ischaemia, increased sympathetic tone, inflammation (systemic as well as myocardial), and electrolyte imbalance. Our finding of a low incidence of myocarditis is consistent with several other reports and a recent review of autopsy cases^31^. Acute respiratory infections, such as influenza, are recognised triggers for adverse CV events in susceptible individuals^32,33^. Additionally, in 277 hospitalised patients with COVID-19 who underwent clinically-indicated echocardiography, biomarker-defined myocardial injury only conferred an increased risk of mortality among individuals with echocardiographic abnormalities^34^. No significant prognostic impact of myocardial injury was observed in patients with structurally normal hearts. These data are consistent with the concept that severe respiratory viral infections may increase susceptibility to further cardiac dysfunction and injury in individuals with pre-existing CVD, and that an ensuing vicious cycle linking cardiac and respiratory dysfunction may drive mortality in COVID-19.

### Limitations

Our analysis was limited to individuals who required hospital admission and are therefore only generalisable to this population. A small minority of patients were still in hospital and were censored at the study end-date (2.1%). However, a sensitivity analysis in patients who were either discharged or died revealed similar findings (**Supplemental Figure 3**). The availability and feasibility of cardiac magnetic resonance imaging, as part of the revised Lake Louise 2018 Criteria^35^ for the diagnosis of myocarditis, was reduced during the pandemic peak, which may have underestimated the incidence of myocarditis. Finally, our multivariable analyses adjusted for the presence or absence of several comorbidities, however measures of control (e.g. blood pressure control for hypertension or HbA1c for diabetes) were not assessed and may impact the risk of death.

## Conclusions

Among patients hospitalised with COVID-19, pre-existing established CVD appears to be a more important contributor to in-hospital mortality than CV risk factors without co-existent CVD, particularly in patients below the age of 70 years. This enhanced risk may be driven, at least in part, by a higher incidence of cardiac complications and myocardial injury in patients with pre-existing CVD whereas VTE appears less important. Optimal management of pre-existing CVD and vigilance for the occurrence of cardiac complications may serve to modify outcomes related to COVID-19 in this group.

## Data Availability

The authors declare that all data supporting the findings of this study are available within the article (and its supplementary information files). Individual participant data will not be made available due to confidentiality regulations.

## Acknowledgements

We are extremely grateful to all the clinicians and NHS staff who have looked after our patients with COVID-19. We additionally thank the patient experts of the KERRI committee.

## Supplemental Materials

### Supplemental methods

*Classification of Cardiovascular Diseases (CVD) and complications of COVID-19* Myocardial infarction, heart failure, myocarditis, pericarditis were defined according to clinician diagnoses in accordance with internationally recognised guidelines^37-40^. Valvular heart disease refers to ‘severe’ grade disease (i.e. not including mild and moderate disease), as defined by European Society of Cardiology guidelines^20^. A pre-existing or new diagnosis of cardiac arrhythmia was defined by the SNOMED CT UK extension parent term “Clinical Arrhythmia” and all daughter terms, and therefore encompasses a wide spectrum of cardiac arrhythmias, including a specific term for atrial fibrillation (which we also report separately). Atrial fibrillation including paroxysmal, persistent and longstanding persistent classifications. Endocarditis was defined according to international guidelines.^41^

Acute myocardial infarction, myocarditis, pericarditis and endocarditis were defined as complications of COVID-19 if a clinician-diagnosis was documented during the hospital admission for COVID-19, consistent with guideline-directed diagnostic criteria^37,39-41^. Acute heart failure complicating COVID-19 was defined as a clinician-made diagnosis of acute left or right ventricular failure e.g. acute pulmonary or peripheral oedema. An exacerbation or decompensation of arrhythmia (including atrial fibrillation) was defined by new or worsening symptoms, haemodynamic instability or poor heart rate control, as documented by the treating physician.

**Supplemental Figure 1.**
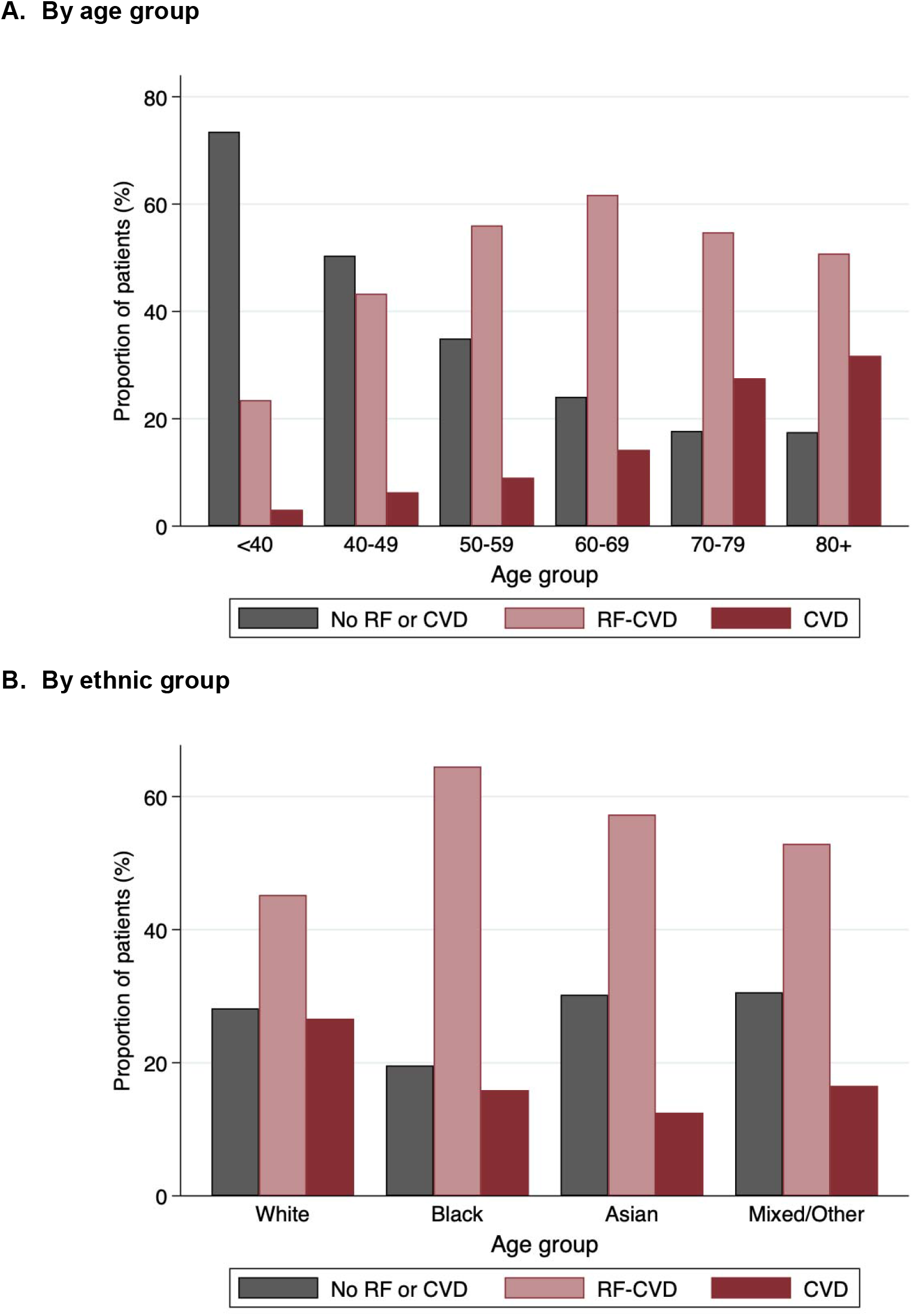
Prevalence of pre-existing cardiovascular disease and risk factors. Bars represent the proportion of patients (%) within each age or ethnic group. CVD, cardiovascular disease; RF, cardiovascular risk factors; RF-CVD, cardiovascular risk factors without established cardiovascular disease;

**Supplemental Figure 2.**
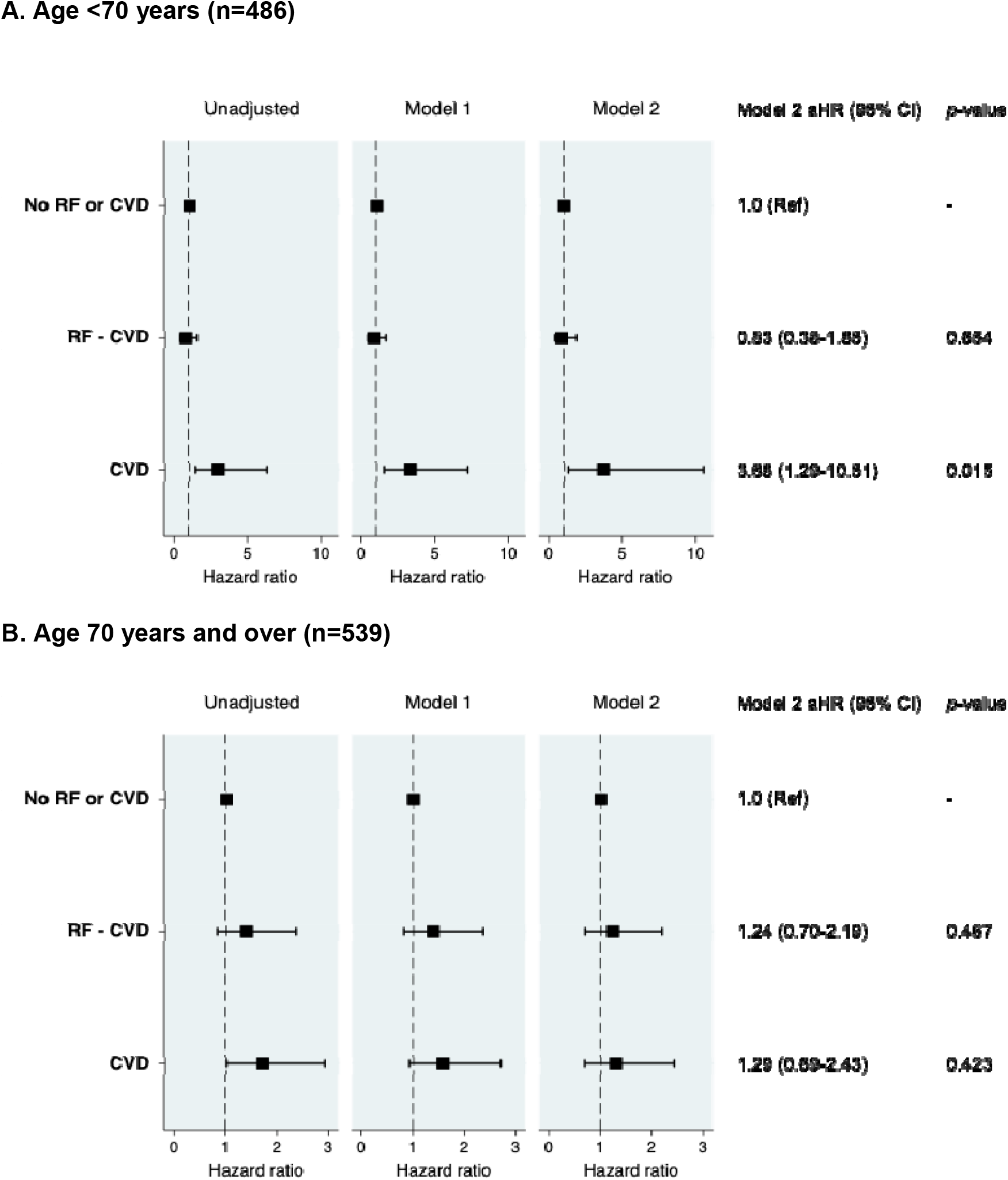
Risk of in-hospital mortality stratified by age and cardiovascular risk group in patients with BMI data available. aHR, adjusted hazard ratio; CVD, cardiovascular disease; RF, cardiovascular risk factors; RF-CVD, cardiovascular risk factors without established CVD. Model 1 adjusted for age, sex and ethnicity. Model 2 adjusted for age, sex, ethnicity, BMI, non-cardiac comorbidities (asthma, COPD, chronic renal failure, pulmonary embolism, DVT) and medications (ACEI or ARB, aldosterone receptor antagonists, beta-blockers, calcium channel blockers, loop diuretics, statins, anticoagulants, antiplatelet agents).

**Supplemental Figure 3.**
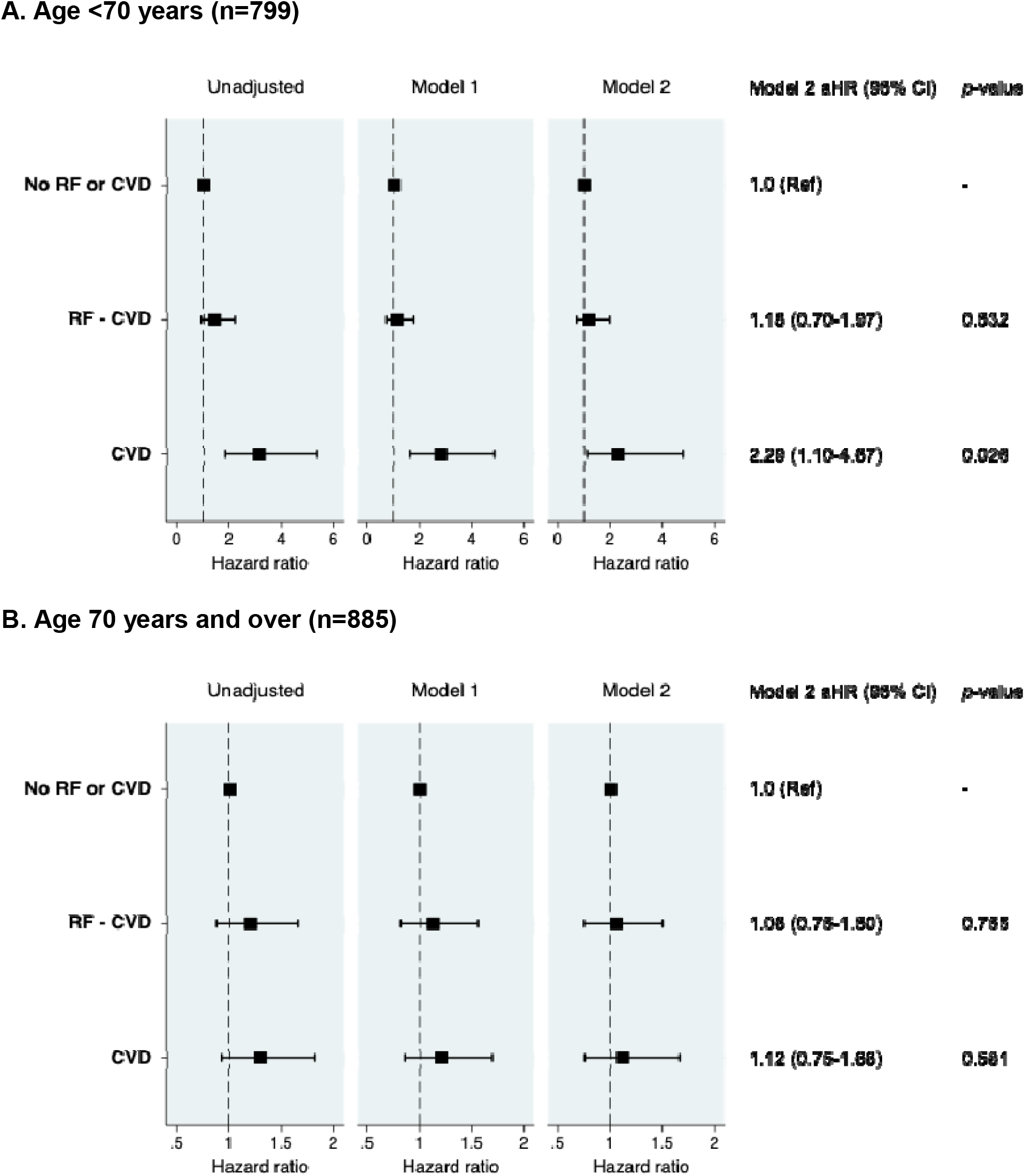
Risk of in-hospital mortality stratified by age and cardiovascular risk group in patients who were discharged or died (i.e. excluding current in-patients). aHR, adjusted hazard ratio; CVD, cardiovascular disease; RF, cardiovascular risk factors; RF-CVD, cardiovascular risk factors without established CVD. Model 1 adjusted for age, sex and ethnicity. Model 2 adjusted for age, sex, ethnicity, BMI, non-cardiac comorbidities (asthma, COPD, chronic renal failure, pulmonary embolism, DVT) and medications (ACEI or ARB, aldosterone receptor antagonists, beta-blockers, calcium channel blockers, loop diuretics, statins, anticoagulants, antiplatelet agents).

**Supplemental Figure 4.**
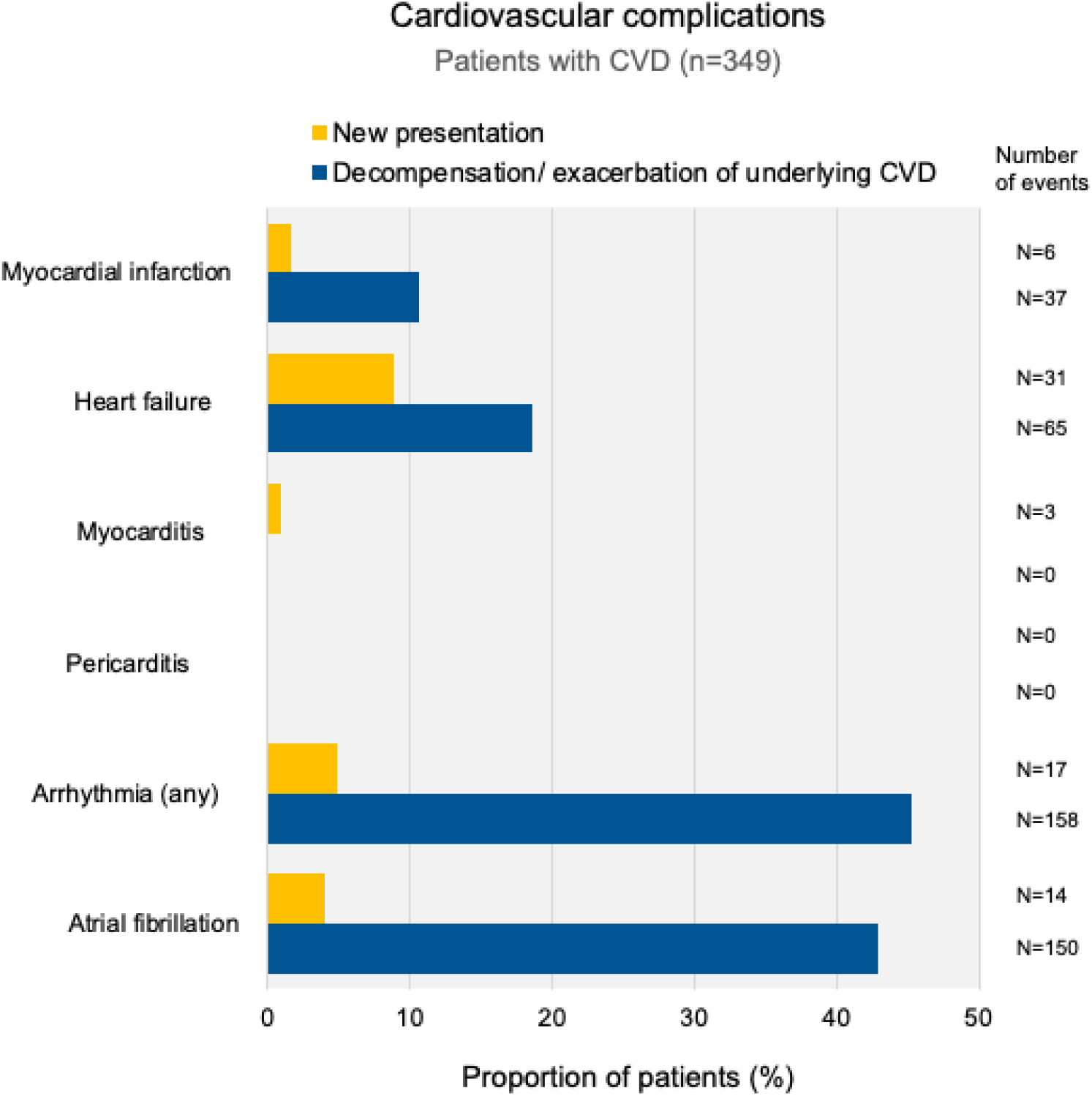
Incidence of COVID-19 related cardiovascular complications in patients with pre-existing established cardiovascular disease. CVD denotes established cardiovascular disease Bars represent the proportion (%) of patients with CVD experiencing that complication. The blue bars represent events occurring in patients with a previous (or underlying chronic) diagnosis of the respective condition, e.g. myocardial infarction occurring in an individual with a previous myocardial infarction, and acute decompensated heart failure occurring in an individual with known chronic heart failure. Exacerbation/decompensation of arrhythmia-related complications (including atrial fibrillation) refer to new or worsening symptoms, haemodynamic instability or poor heart rate control. Note that patients can experience more than 1 complication. Total number of cases displayed at the end of each bar.

**Supplemental Figure 5.**
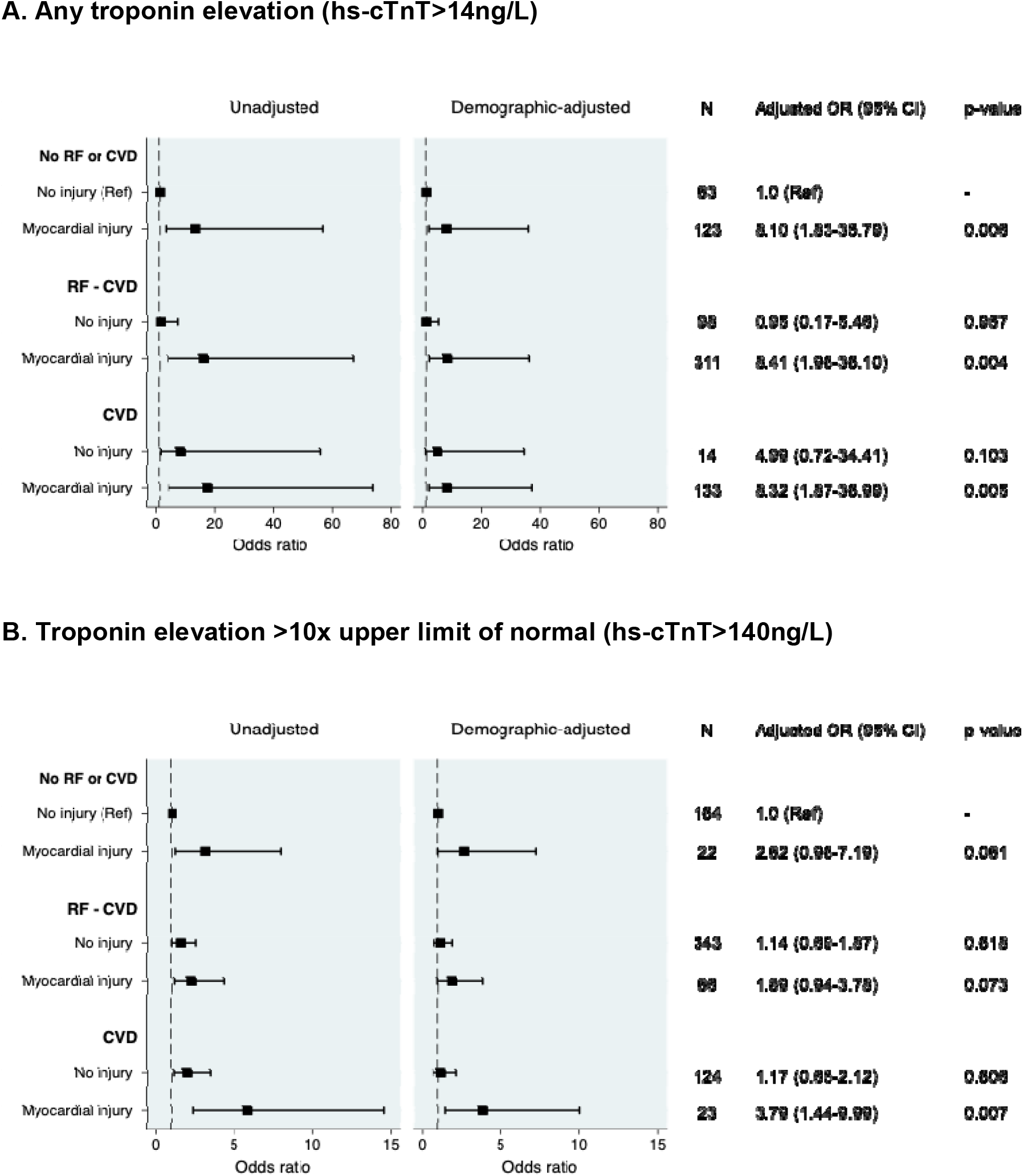
Risk of in-hospital mortality stratified by age, cardiovascular risk group and myocardial injury (n=742 patients) aOR, adjusted odds ratio; CVD, cardiovascular disease; RF, cardiovascular risk factors; RF-CVD, cardiovascular risk factors without established CVD. Demographic-adjusted model: adjusted for age, sex and ethnicity.

